# Single-cell RNA sequencing identifies aberrant transcriptional profiles of cellular populations and altered alveolar niche signalling networks in Chronic Obstructive Pulmonary Disease (COPD)

**DOI:** 10.1101/2020.09.13.20193417

**Authors:** M Sauler, JE McDonough, TS Adams, N Kothapalli, JC Schupp, J Nouws, M Chioccioli, N Omote, C Cosme, S Poli, EA Ayaub, SG Chu, KH Jensen, J Gomez-Villalobos, CJ Britto, MSB Raredon, PN Timshel, N Kaminski, IO Rosas

## Abstract

Chronic Obstructive Pulmonary Disease (COPD) pathogenesis involves a failure to maintain alveolar homeostasis due to repetitive injury and inflammation. In order to improve our understanding of cell-specific mechanisms contributing to COPD pathogenesis, we analysed single-cell RNA sequencing (scRNAseq) profiles of explanted parenchymal lung tissue from 17 subjects with advanced COPD requiring transplant and 15 control donor lungs. We identified a subpopulation of alveolar type II epithelial cells that uniquely express *HHIP* and have aberrant stress tolerance profiles in COPD. Amongst endothelial cells, we identified overlapping and unique shifts in transcriptional profiles of endothelial subtypes that may contribute to vascular inflammation and susceptibility to injury. We also identified population composition changes amongst alveolar macrophages. Finally, application of integrative analyses to our scRNAseq data identified cell-specific contributions to COPD heritability and dysfunctional cell-cell communication pathways that occur within the COPD alveolar niche. These findings provide cell type-specific resolution of transcriptional changes associated with advanced COPD that may underlie disease pathogenesis.

## INTRODUCTION

Chronic Obstructive Pulmonary Disease (COPD) is characterized by persistent airflow obstruction and the pathologic findings of small airway inflammation and parenchymal tissue destruction (i.e. emphysema).^1,2^ While COPD is commonly caused by aerosolized pollutants such as cigarette smoke (CS), the clinical and biologic effects of CS are heterogenous. Therefore, complex interactions of genetic factors, host responses, and environmental exposures underlie COPD pathogenesis.

Previous studies of COPD pathogenesis based on murine models of disease, genetic association studies, or analyses of bulk human lung tissue have greatly expanded our understanding of disease pathogenesis.^3-5^ The pathogenic cascade of COPD is initiated by repetitive insults to epithelial and endothelial cells within the distal airways and alveolar niche. This engenders activation of diverse cellular processes including immune cell infiltration, extracellular matrix proteolysis, cellular metabolic dysfunction, loss of proteostasis, DNA damage, autophagy, cellular senescence, and activation of regulated cell death pathways.^4,6,7^ Consequently, there is an inability to maintain alveolar homeostasis, chronic inflammation, and alveolar septal destruction, particularly in advanced COPD.^4^ However there remains limited knowledge of cell type-specific mechanisms or complex interactions amongst the multiple lung cell types that contribute to disease progression.

Recent studies have used single-cell RNA sequencing (scRNAseq) to obtain gene expression profiles at single-cell resolution and identify novel cellular phenotypes and disease mechanisms.^8-12^ Herein, we analysed scRNAseq profiles of parenchymal tissue obtained from explanted lungs of patients with advanced COPD requiring lung transplant and control donor lungs. Through this approach, we characterized aberrant transcriptional profiles of epithelial, endothelial, and immune cell populations. We also integrated genome-wide association study (GWAS) data with our scRNAseq analysis to determine cell types contributing to COPD heritability and applied ligand-receptor connectome analyses to identify dysregulated cell-cell interactions. These findings suggest novel cell-type-specific mechanisms that may contribute to disease pathogenesis.

## METHODS

### Sample collection and single-cell RNA sequencing

Tissue procurement, sample processing and single-cell sequencing was previously performed, and methods are further detailed in supplemental methods.^13^ Briefly, scRNAseq was performed on parenchymal lung tissue, either from subjects with advanced COPD undergoing lung transplantation or from rejected donor lungs without disease that were used as controls. Study protocols were approved by Partners Healthcare Institutional Board Review (IRB Protocol 2011P002419).

### Clustering, differential cell expression and regulon analysis

Clustering and differential expression was performed using the Seurat package (v.3.2.0) in R. Overall marker genes for each cell type were identified by calculating the diagnostics odds ratio (DOR) for each gene, and cell types were annotated using canonical marker genes as previously described.^13^ We identified regulons (i.e. imputed transcription factors predicted to regulate gene expression) using pySCENIC (v.0.10.2).^14^ Further details are available in supplemental methods.

### CELL-type Expression-specific integration for Complex Traits (CELLECT)

We used CELLECT to quantify associations between cell-type specificity of expressed genes and findings from genome-wide association studies (GWAS) of lung function and COPD.^15,16^ CELLECT has been previously described^17^ and furthered detailed in supplemental methods. Briefly, CELLECT generates a genetic prioritization scores for each gene based on cell type-expression specificity and GWAS summary statistics for related genetic variants. It then prioritizes cell types enriched for genes with high genetic prioritization scores. As input to CELLECT, we used summary statistics derived from genetic studies of UK Biobank data for presence of COPD^15^ and lung function.^16^

### Lung connectome to identify cell-cell interactions

Methods used to generate the lung connectome have been previously described and are detailed in supplemental methods.^18^ Briefly, average expression values for every gene within cell types were calculated and mapped using the FANTOM5 database of known ligand-receptor pairs. A node was defined as a cell type. An edge connecting two nodes was generated if > 5% of the cells within each node expressed the cognate molecule of the ligand-receptor pair, and edge-weights represented the product of the average expression values of the ligand and receptor. For the alveolar niche network, both non-directional and directional (i.e. incoming and outgoing) cumulative edge-weights were calculated using the sum of edge-weights computed from the average scaled expression values of all edges between two nodes. For the pathway centrality analysis, we only used ligand-receptor pairs preassigned to specific signaling modes (i.e. signaling pathways). Cumulative out- and incoming edge-weights were calculated using average unscaled expression values computed for each individual signalling mode. For these analyses, we also calculated Kleinberg hub and authority scores which are metrics of outgoing and incoming centrality that take into account the number of edges and edge-weights connecting a node within a network.

## RESULTS

We analysed scRNAseq profiles of explanted lung tissue parenchyma obtained from 17 patients with advanced COPD, and 15 age-matched donor lungs as controls (**Figure 1A**). Demographics and pulmonary function test results are shown in **Supplemental Table 1**. There were eight females in both groups and the median age of all subjects was 62 years old (range 41–80). All advanced COPD subjects had radiographic evidence of emphysema. COPD subjects were former smokers; four of the donors were either current or former smokers. The final dataset consisted of 49,976 cells from control donor lungs and 61,564 cells from COPD lungs, and we identified 37 distinct cell types in both control and COPD lungs based on representative marker genes (**Figures 1B**). Canonical markers of identified cell types are shown. (**Figure 1C–E)**. Data has been deposited in the Gene Expression Omnibus (GSE136831) and can be explored using our online portal (www.copdcellatlas.com).

**Figure 1.**
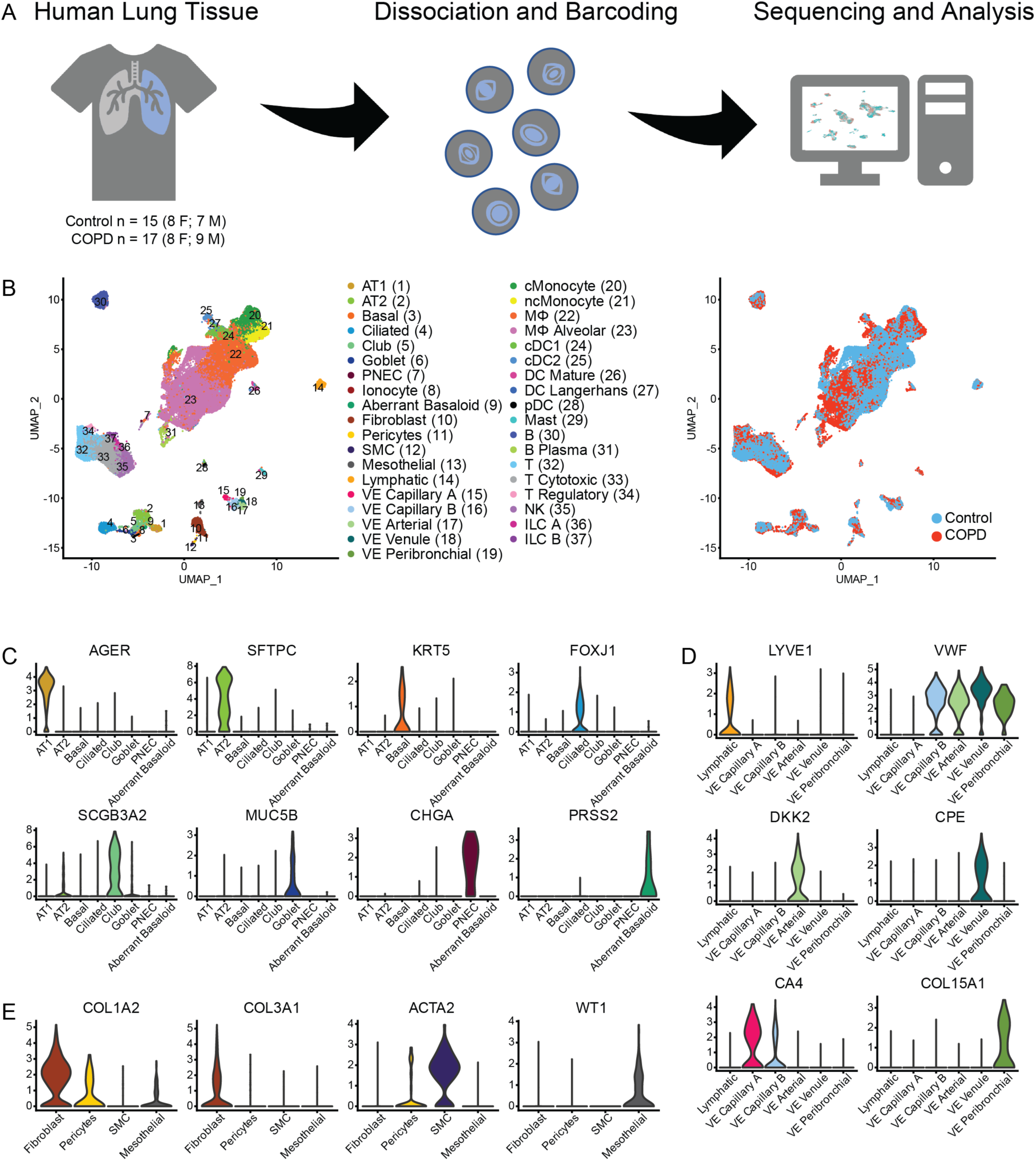
Profiling of cell types in COPD using scRNAseq data. (**A**) Overview of project design. Tissue from 17 lungs with advanced COPD and 15 control donor lungs were dissociated into single cell suspensions. Individual cells were barcoded and sequenced for analysis. (**B**) Uniform Manifold Approximation and Projection (UMAP) representation of 111,540 single cells grouped into 37 distinct cell types (left) with identification of COPD and control cells (right). Violin plots of normalized expression values for canonical cell-specific marker genes for (**C**) epithelial, (**D**) endothelial, and (**E**) stromal cells.

### Epithelial cells from advanced COPD lungs exhibit transcriptional evidence of aberrant differentiation and impaired stress responses

We identified all major epithelial cell types including alveolar type I (AT1) and alveolar type II (AT2) cells (**Figure 2A**). Amongst all epithelial cells, there were no significant cell proportion differences between COPD and control lung tissue except for a small population of 33 aberrant basaloid cells, from 9 COPD subjects (range: 1–10 cells per subject), which we recently identified in IPF (**Figure 2B**).^13^ These cells do not express basal markers *KRT5* and *KRT15* but do express *TP63, KRT17, LAMC2*, and senescence-related genes (*CDKN1A, CDKN2A*, *GDF15)*.

Two distinct sub-clusters of AT2 cells (AT2_A_ and AT2_B_) were observed in COPD and control donor lungs (**Figure 2C**). While both clusters expressed AT2 markers such as *SFTPC* and *SFPTA1*, the most striking difference between the two clusters was a markedly greater expression of these genes in the AT2_B_ population. AT2_B_ cells also expressed higher levels of tissue protease *PGC* and the chemokine *CXCL2*, and almost uniquely express *HHIP*, a common GWAS association for COPD and emphysema.^19^ Regulon analysis of imputed transcription factors suggested increased activity of *ETV1* and *ETV5*, which are involved in maintaining AT2 cell identity.^20^ In contrast, AT2_A_ cells demonstrated a relative increase in the expression of genes implicated in AT2 stem cell function including *CDH1*,^21^ WNT pathway gene *TNIK*,^22^ tyrosine kinase receptor *ERBB4*,^23^ and decreased expression of the WNT inhibitory protein *WIF1*. Regulon analysis suggested increased activity of transcription factors that potentiate or mediate HIPPO/YAP and WNT signalling pathways including *FOXP1*, *FOXP2*, *TEAD1/3/4*, *NFATC2*, and *STAT3*. Therefore, AT2_A_ resemble WNT-responsive AT2 epithelial progenitor cells (AEPs) and AT2_B_ resemble non-AEP AT2 cells that express higher levels of canonical AT2 cell markers.^24-26^

Amongst all epithelial cells, AT2_B_ cells had the greatest number of differentially expressed genes (DEGs) between control and COPD lung tissue, followed by ciliated, AT1, and AT2_A_ cells (**Supplemental table 2**). AT2_B_ cells from COPD lungs had decreased expression of genes involved in mitigating oxidative and toxin-mediated stress (*MGST*, *AKR1A1*, *TXND17*, *GSTP1*, *ALDH2*) and genes encoding S100 family proteins (**Figure 2D**). The most significantly decreased expressed gene was *NUPR1* (fold change (fc)=0·35; adj. P=2·75×10^-17^), a stress-induced protein that orchestrates different aspects of the cellular stress response.^27^ In AT2_A_ cells, we identified decreased gene expression and regulon activity related to cytoskeletal rearrangement (*KAZN, ANK3, UTRN*, *KANK1*) and epithelial differentiation (*NHSL1, ELF3, GPC6, MEF2D*) (**Figure 2E**). Together, these data highlight transcriptional profile shifts that may contribute to impaired AT2_B_ cellular stress responses and impaired AT2_A_ epithelial renewal with advanced COPD.

**Figure 2.**
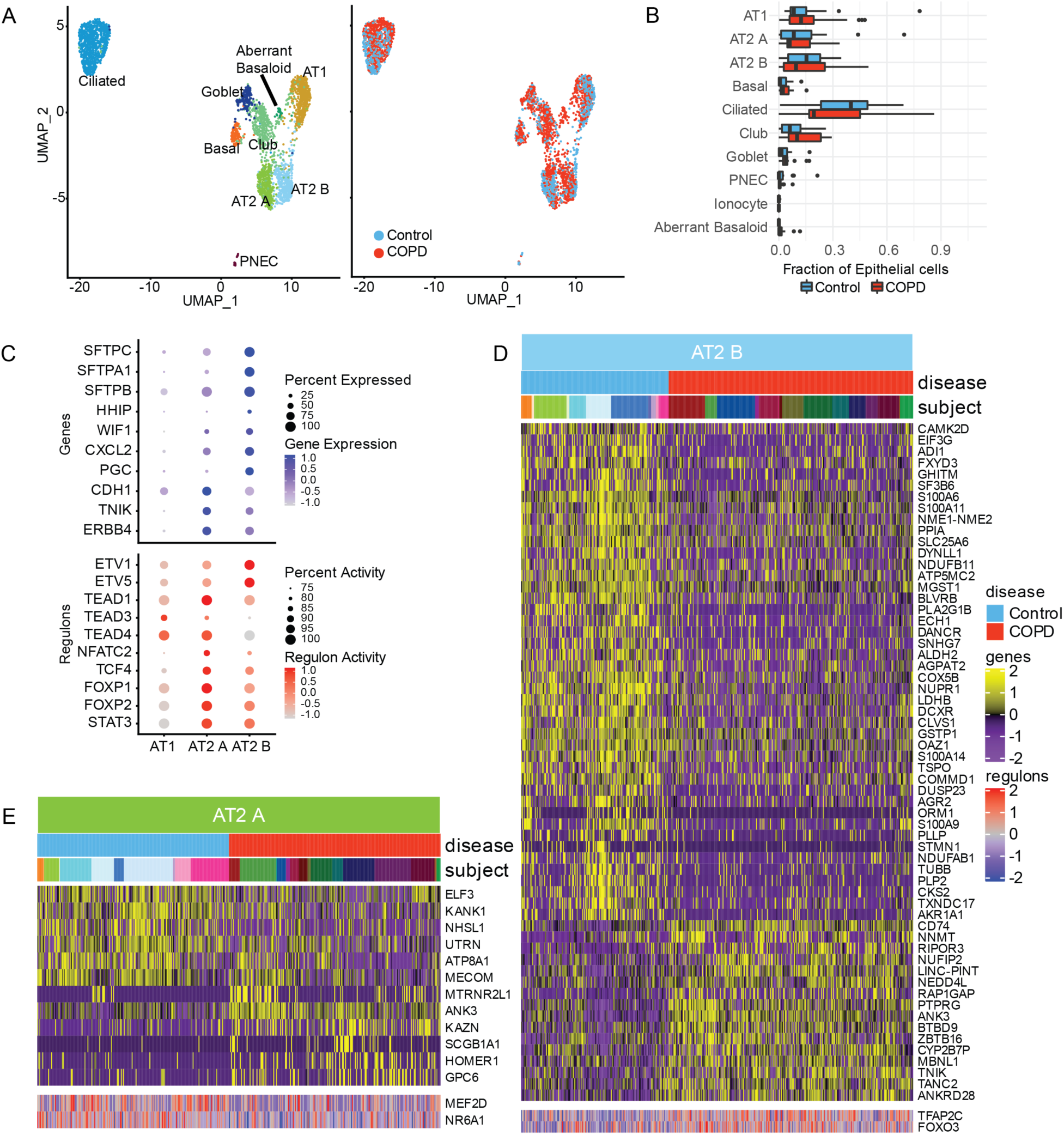
Epithelial cells in COPD demonstrate dysfunction in AT2 subtypes. (**A**) UMAP of epithelial cells from COPD and control donor lungs. Samples are colour labelled by cell type (*left*) and disease category (*right*). Alveolar type II (AT2) cells can be distinguished as two clusters denoted as AT2_A_ and AT2_B_. (**B**) Distribution of subject-specific epithelial cell types as a fraction of the total number of epithelial cells per disease category. Boxes represent interquartile ranges (IQRs); whiskers are 1·5 x IQR; dots represent subjects (**C**) Dot plot of z-scores for marker gene expression values (*top*) and predicted transcription factor (i.e. regulon) activity (*bottom*) of AT2_A_ and AT2_B_ cells. Dot size reflects percentage of cells with gene expression or regulon activity; colour corresponds to degree of expression (gene) or activity (regulon). (**D, E**) Heatmaps of z-scores for differentially expressed genes (yellow/purple) and predicted transcription factor (i.e. regulon) activity (red/blue) between control and COPD for AT2_B_ and AT2_A_ cells (Wilcoxon rank sum test, FDR <0.05). Each column represents expression values for an individual cell. Columns are hierarchically ordered by disease phenotype and subject, in which disease category and individual subject are represented by unique colours. z-scores were calculated across all epithelial cells.

### COPD-specific features of endothelial cells

Endothelial injury is implicated in COPD pathogenesis, but the specific changes in the human lung are not thoroughly understood. Clustering identified two distinct populations of capillary cells (denoted as capillary A and B), as well as arterial, venous, lymphatic, and systemic peri-bronchial endothelial cells (**Figure 3A**). No significant differences in the proportions of endothelial cells between disease and control were detected (**Supplemental Figure 1**). We compared DEGs between COPD and control cells for each endothelial cell type independently. While the majority of DEGs were unique to each cell type, we observed that many DEGs overlapped across multiple endothelial cell types. (**Figure 3B**). Notably, 30 genes were differentially expressed across three or more endothelial cell types, including increased expression of metallothionines (*MT1X, MT1M, MT2A, MT1E*) and AP-1 subunits (*FOS, FOSB, JUND*), and decreased expression of genes related to angiogenesis (*ID1, ID3, LDB2*) (**Figure 3C**). The largest set of overlapping endothelial DEGs occurred between both capillary cell types and involved 20 DEGs demonstrating increased expression of genes involved in inflammatory signalling (*CX3CL1, IL32*), cellular stress responses (*GADD45B*), and vesicular trafficking (*WASHC2C, WASHC2A*), and decreased expression of genes that can promote endothelial repair (*SEMA6A*, *WNT2B*).^28,29^ Capillaries also exhibited cell type-specific increases in the expression of inflammatory genes such as *TNFRSF10D, ITPKC*, and *IRF1* in capillary A and *TNFAIP3, IFI5*, and *IL6* in capillary B, as well as cell type-specific decreases in the expression of genes that promote endothelial angiogenesis and homeostasis such as *HIF3 and MEF2C* in capillary A and *TFP1* in capillary B.^30^ These data demonstrate overlapping and distinct DEGs amongst lung endothelial cell types that may contribute to pathologic endothelial activation or impaired endothelial repair in COPD.

**Figure 3.**
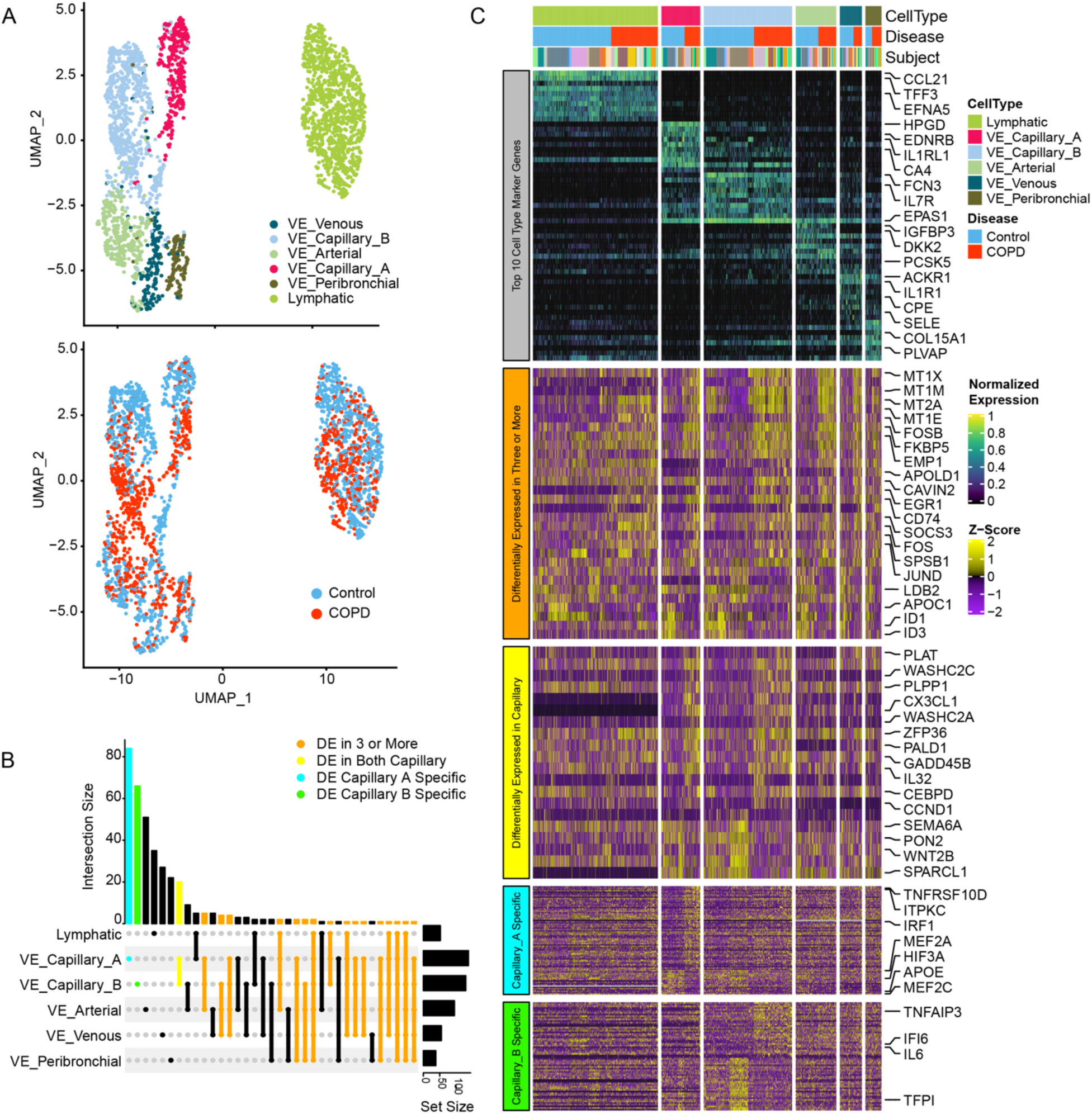
COPD endothelial cell types demonstrate universal and cell type-specific transcriptional aberrations. (**A**) UMAPs of all vascular endothelial (VE) and lymphatic endothelial cells from control and COPD subjects. UMAPs are colour labelled by cell type (*top*) and disease status (*bottom*). UpSet plot visualizing the properties of intersecting and unique sets of differentially expressed (DE) genes between COPD and control amongst endothelial (Wilcoxon rank sum test, p < 0.001, fc > 0.5). (**C**) Heatmap of corresponding differentially expressed genes between COPD and control amongst six subtypes of endothelial cells. Each column represents expression values for an individual cell. Columns are hierarchically ordered by endothelial subtype, disease phenotype, and then subject. Grey row (*top)*: expression values for top marker genes are unity normalized between 0 and 1 across all endothelial subtypes. Orange row (*middle*): z-scores of differentially expressed genes in three or more endothelial cell types between control and COPD. Yellow row (*middle*): z-scores of differentially expressed genes in both capillary A and capillary B cells between control and COPD. Blue and green row (*bottom*): z-scores of differentially expressed genes unique to capillary A (blue) or capillary B (green). Unity normalization and z-score calculations were performed using all endothelial subtypes.

### Transcriptional heterogeneity of alveolar macrophages

We explored the immunologic landscape of the distal lung and identified 18 distinct immune cells. (**Supplemental Figure 2**) Between control and COPD, we observed significant differences in cell compositions amongst interstitial macrophages (fc=0·48; P=4·5×10^-4^), non-classical monocytes (fc=2·18; P=0·026), plasmacytoid dendritic cells (fc=3·46; P=8·1×10^-4^), conventional dendritic cells (fc=2·79; P=0·010), mature dendritic cells (fc=1·89; P=0·046), and mast cells (fc=7·70; P= 4·5×10^-4^) (**Supplemental Figure 1**). The largest population of cells were alveolar macrophages which have been implicated in alveolar inflammation and tissue destruction.^31,32^ To characterize the heterogeneity of alveolar macrophages, we re-embedded the alveolar macrophage cluster in UMAP space and re-clustered these cells into eight clusters (**Figure 4A**). We then compared the relative abundance of the eight clusters between control and COPD and identified changes in alveolar macrophage population composition between COPD and control; cluster-0 macrophages were enriched amongst controls while cluster-5 macrophages were enriched amongst patients with COPD (adjusted p <0.05) (**Figure 4B**). The corresponding cluster markers for cluster-5 and cluster-0 are shown in **Figure 4C**. Cluster-0 markers included heat shock proteins (*HSPA1A, HSPA1B, HSPH1, HSPD1, HSP90AA1, HSPE1*), while the top cluster-5 markers were metallothionines (*MT1G, MT1X, MT1E, MT2A, MT1M, MT1F, MT1H, MT1A, MT1L*). There were also multiple DEGs between COPD and control macrophages that were not represented in any specific cluster. The most significant DEGs included genes associated with macrophages chemotaxis and inflammation (*THBS1, PELI1*, and *CDC42)* (**Figure 4D**). These results highlight changes in macrophage population composition as well as common DEGs across the multiple macrophages subpopulations.

**Figure 4.**
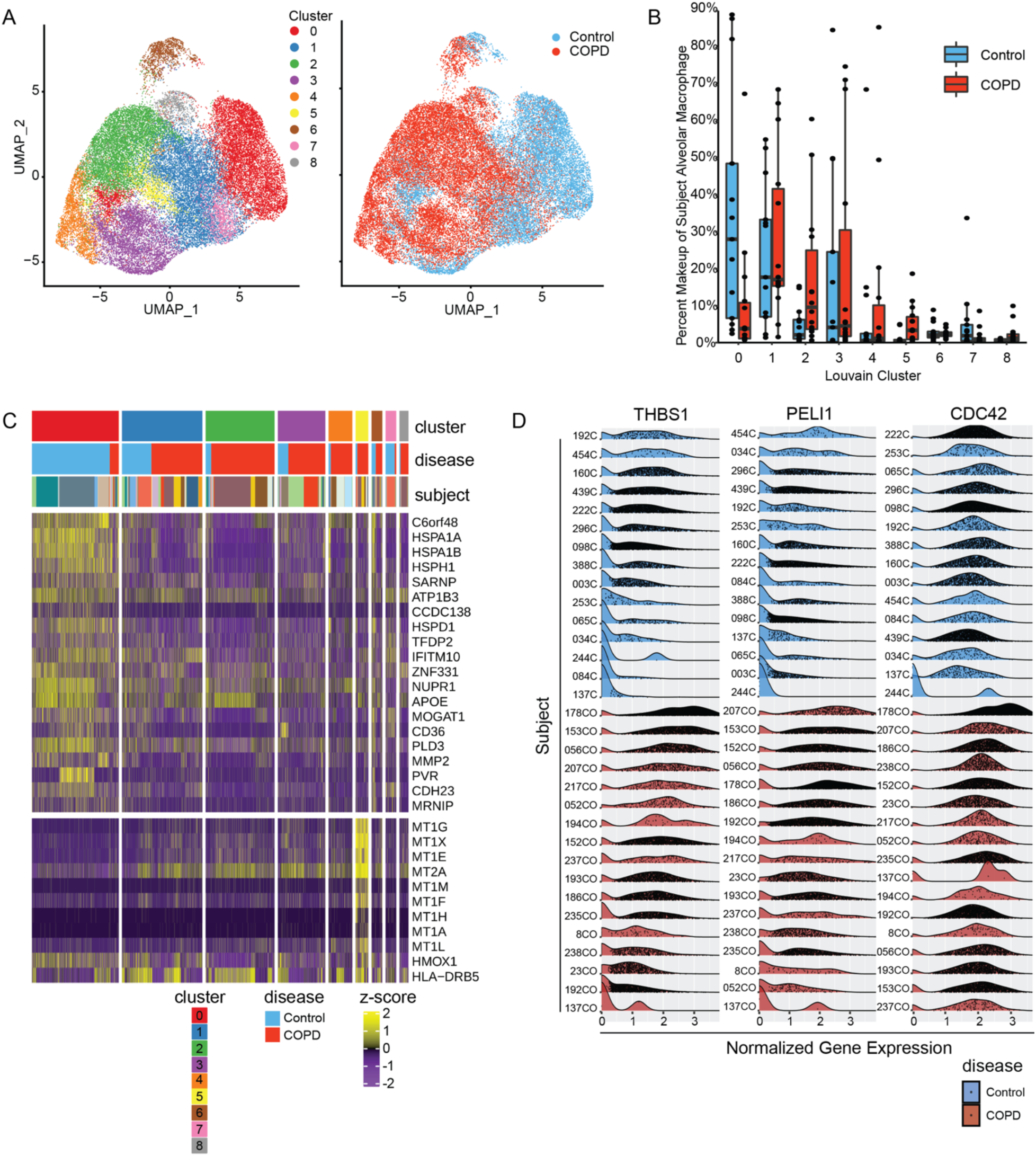
Changes in alveolar macrophage population composition in COPD. **(A)** UMAPs of control and COPD alveolar macrophage cells, colour labelled by Louvain cluster *(left)* and disease status (right). **(B)** Percent makeup of alveolar macrophage across all nine Louvain clusters per subject, grouped by disease state. Boxes represent interquartile ranges (IQRs); whiskers are 15 × IQR; dots represent subjects. The percent of cluster 0 and cluster 5 alveolar macrophages are different between control and COPD (Wilcoxon rank sum test, FDR <0.05). **(C)** Heatmap of the distribution of z-scores of marker genes for cluster 0 and cluster 5 alveolar macrophages. Columns represent expression values from individual cells and are hierarchically ordered by macrophages cluster, disease status, and subject. **(D)** Density plots of *THBS1, PELI1* and *CDC42* gene expression across all alveolar macrophage cells, grouped by control (blue) and COPD (red) subjects. Within each disease group, subjects are ordered top to bottom from highest within-disease group expression average to lowest.

### COPD genetic variants enrich for alveolar niche cells

The cell types through which genetic variants mediate COPD susceptibility are not well-defined.^33^ We sought to address this by using CELLECT, a computational tool that colocalizes scRNAseq and GWAS data. Using this approach, we identified multiple cell types enriched for GWAS-associated genes including AT2, smooth muscle, and fibroblast populations (**Figure 5A**). Notably, we did not identify immune cell populations with high prioritization scores despite adequate representation of immune cells in our dataset. For reference, the top genes commonly associated with COPD with genetic prioritization scores > 0.5 are shown (**Figure 5B**). Collectively, these data suggest that much of COPD heritability is mediated by non-hematopoietic lung resident cells.

**Figure 5.**
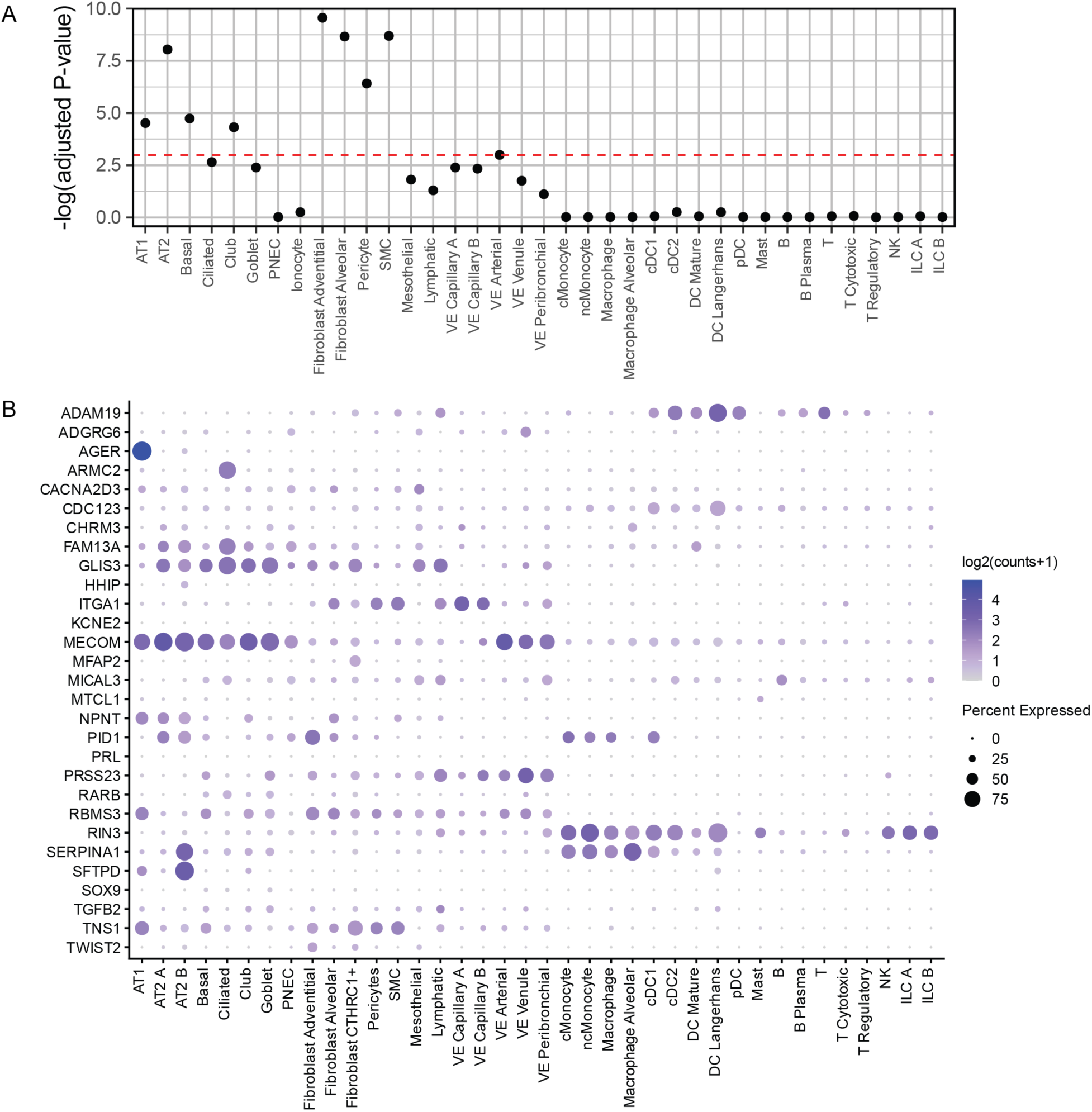
CELLECT analysis identifies cellular basis for COPD heritability. **(A)** Plot of negative log adjusted p-values for cell type-specific enrichment for GWAS-identified genes associated with lung function. Red line denotes threshold of significance (FDR < 0.05). Data showing high enrichment scores for non-hematopoietic cells are associated with COPD heritability traits. **(B)** Dot plot of z-scores for commonly associated GWAS genes that were highly ranked in CELLECT analysis. Dot size reflects percentage of cells with gene expression; colour corresponds to degree of expression.

### Alveolar niche network and pathway centrality analysis reveal changes in alveolar signalling topology

We performed connectome analyses to characterize alveolar signalling topologies and identify differences in cell-cell communication between control and COPD lungs. We generated network-level maps based on predicted ligand-receptor interactions amongst alveolar cell types (**Figure 6A**). To quantify changes in cell-cell interactions, we measured differences in directed cumulative edge-weights (i.e. cumulative outgoing or cumulative incoming edges) between control and COPD (**Figure 6B**). In COPD, there were increases in directed edge-weights between nodes of capillaries, monocytes, and macrophage, with the greatest increase in edge-weight occurring between non-classical monocytes and capillaries. We then sought to determine the contribution of individual signalling pathways to COPD-related changes in alveolar signalling topology. Ligand-receptor interactions were preassigned to canonical signalling pathways and centrality metrics were calculated, including incoming and outgoing edge-weights and Kleinberg hub and authority scores. These centrality rankings prioritize specific cell types responsible for cell-cell interactions through these individual pathways. We found changes in 16 out of 22 signalling pathways, including inflammatory pathways (e.g. CC-motif chemokines, CXCL-motif chemokines, interleukins, and TNF) and canonical signalling pathways previously implicated in COPD pathogenesis (VEGF, PDGF, MET, WNT and TGF-*β*) (**Figure 6C**). We did not identify changes in FGF, Ephrin, and NOTCH signalling (**Supplemental Figure 3**). To quantify changes in cell-cell interactions, we generated individualized pathway centrality networks for each subject, and compared edge-weights for each signalling mode across subjects between control and COPD. The pathway with the most significant differences in cumulative edge-weights was CXCL-motif chemokine signalling. There was an increase in outgoing CXCL-motif edge-weights from adventitial fibroblasts (fc=2·4; P=2·0×10^-4^) and capillary B cells (fc=4·2; P=3·1×10^-4^), and an increase in incoming CXCL-motif edge-weights towards alveolar macrophages (fc=3·2; P=7·2×10^-4^), interstitial macrophages (fc=3·0; P=4·6×10^-3^), AT2_B_ cells (fc=3·6; P=4·0×10^-3^), and AT1 cells (fc=4·2; P=3·5×10^-2^). (**Figure 6D**).^14^ Collectively, these findings suggest that increased interactions between capillaries, monocytes, and macrophages may be major drivers of alveolar inflammation in COPD, and demonstrate potential pathway-specific changes in cell-cell communication, particularly increases in CXCL-chemokine signalling, occurring within the COPD alveolar niche.

**Figure 6.**
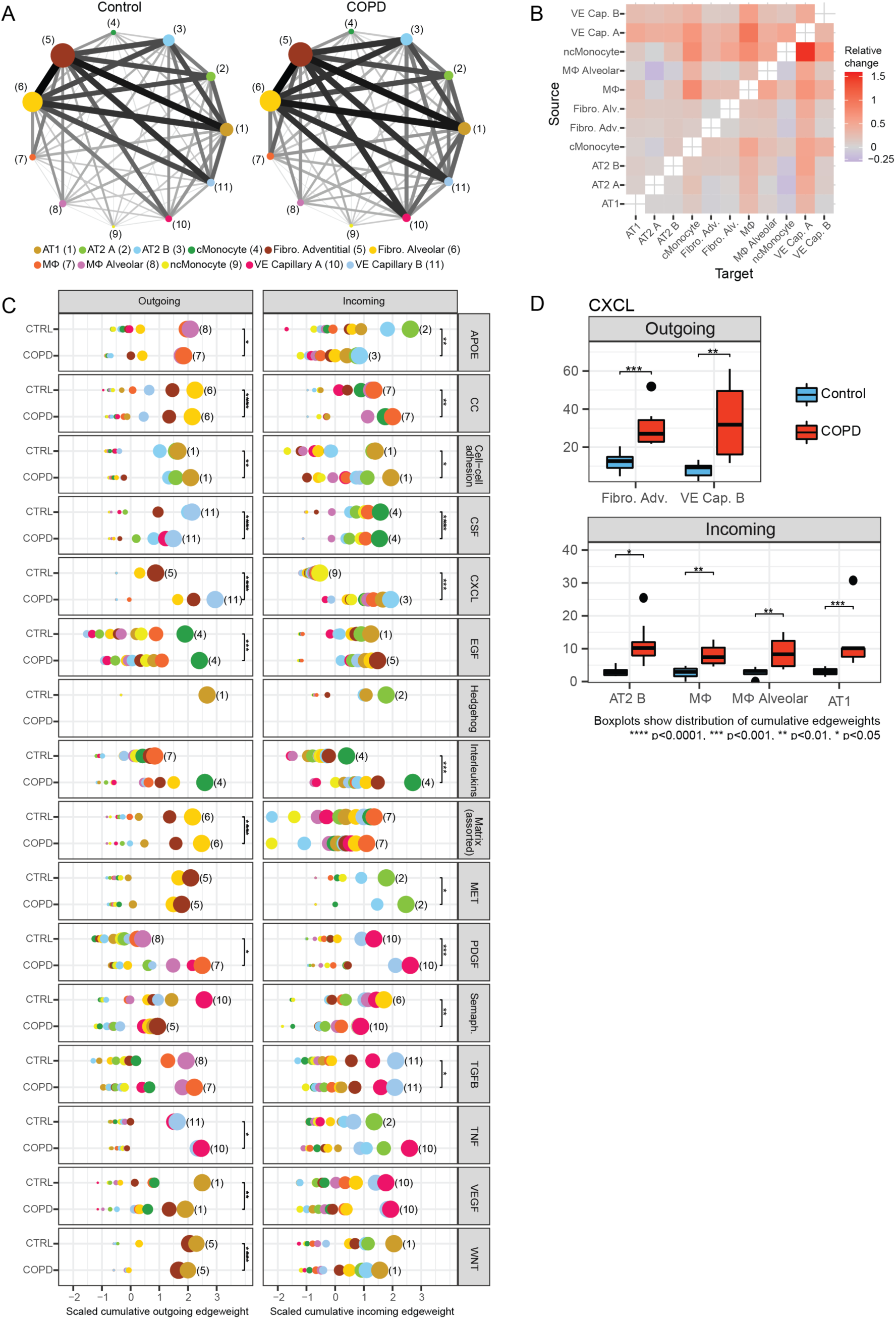
Alveolar niche networks and pathway centrality analyses. **(A)** Network plots of the alveolar niche in control *(left)* and COPD (right). Nodes represent cell types. Node size is proportional to the Kleinberg hub score, and the thickness of each edge is proportional to the sum of all edge-weights (non-directed) between two nodes. Individual cell types are labelled by colour and number. **(B)** Heatmap shows relative changes between control and COPD in cumulative directed edge-weights between outgoing (source) and incoming (target) nodes. **(C)** Centrality analysis comparing the alveolar connectome for each mode of signalling between control and COPD. Panel shows outgoing edge-weights and Kleinberg hub scores *(left)* and incoming edge-weights and Kleinberg authority scores *(right)*. Cumulative edge-weights *(x-axis)* have been scaled for each mode of signalling. Dot size is proportional to the Kleinberg scores for each cell type within a given mode of signalling. Individual cell types are colour labelled as in Figure 6A, and numbers shown identify cell types with the largest cumulative edge-weights. P-values computed using the Durbin test to compare control and COPD across cell types for each signalling mode. **(D)** Box and whisker plots of cumulative edge-weights were computed using patient-level connectomes for CXCL-motif signalling. P-values computed using Wilcoxon rank sum tests to compare cumulative edge-weights between control and COPD subjects. AT1=alveolar type I epithelial cells, AT2=alveolar type II epithelial cells, cMonocyte=classical monocytes, ncMonocyte=non-classical monocytes, VE=vascular endothelial, Fibro=fibroblast, Semaph=semaphorins. MØ=interstitial macrophages, MØ alveolar=alveolar macrophage.

## DISCUSSION

In this study, we leveraged the cell-specific resolution of scRNAseq to improve our understanding of aberrant transcriptional changes in the lung parenchyma that occur with advanced COPD. We identified two subpopulations of alveolar type II epithelial cells, an AT2_B_ population that expresses *HHIP* with transcriptional changes suggestive of impaired stress tolerance profiles and an AT2_A_ subpopulation with transcriptional changes suggestive of impaired epithelial differentiation. We identified augmented stress and inflammatory expression profiles in lung endothelial cells in emphysema and distinct macrophage subpopulations enriched in control and emphysematous lung tissue. Integration of GWAS with our scRNAseq dataset identified lung resident cells as the primary drivers of COPD heritability, while our connectome analysis revealed striking differences in cell-cell interactions, particularly CXCL-chemokine signalling, within the alveolar niche between control and COPD. Together, these findings suggest cell-specific molecular changes that may underlie COPD pathogenesis.

COPD pathogenesis is associated with a failure of AT2 cells to maintain alveolar homeostasis. Previous studies have demonstrated two unique populations of AT2 cells: AEPs are AT2 cells that can function as facultative progenitor cells to regenerate alveolar epithelium and non-AEPs express higher levels of canonical AT2 cell markers.^24-26^ Similarly, we identified two AT2 populations in both control and COPD lungs that correspond to these populations; AT2_A_ cells have a small relative increase in the expression WNT-responsive genes and regulons while AT2_B_ cells have higher expression of surfactant proteins, *HHIP*, and other AT2 cellular markers. Interestingly, the largest difference between AT2_A_ and AT2_B_ cells was the expression of genes related to surfactant production and other homeostatic AT2 functions, suggesting that the contrasting properties of these two cell populations may extend beyond the eponymous functions of AEPs and non-AEPs.

Both AT2_A_ and AT2_B_ populations have divergent transcriptional changes in advanced COPD. The largest number of DEGs was surprisingly identified amongst AT2_B_ cells, that have decreased expression of proteins involved in mitigating oxidative and toxin-mediated stress responses. Additionally, we found that AT2_B_ cells in both COPD and control lungs almost uniquely express *HHIP*, a gene repeatedly associated with COPD and emphysema in genetic association studies.^19^ Previous studies showed haploinsufficient *Hhip* mice are prone to emphysema in part through their susceptibility to oxidative stress,^34^ with the caveat that the cellular distribution of *HHIP* is different between mice and humans. Collectively, these findings suggest unique roles for AT2_A_ and AT2_B_ cell populations in COPD pathogenesis, in which AT2_B_ cells have a diminished capacity to mitigate cellular stress while AT2_A_ cells have an impaired capacity to mediate repair.

Our analysis provides a comprehensive overview of transcriptional changes amongst endothelial cells in advanced COPD.^35-40^ We found evidence for endothelial activation (i.e. pro-inflammatory, pro-coagulant phenotype) as demonstrated by increased endothelial expression of AP1 transcription factor, inflammatory cytokines, cellular stress response genes, and pro-coagulant signalling genes. We also found evidence for a diminished capacity for endothelial repair as suggested by downregulation of genes associated with angiogenesis. The causes of these transcriptional changes may be a direct consequence of oxidative stress from environmental exposures. However, our connectome analyses suggest the possibility that increased crosstalk between capillaries and myeloid-derived cells may also important contributors to endothelial transcriptional changes in advanced COPD. Using our connectome analyses, we identified dysregulated alveolar signalling topologies and prioritized cell types responsible for canonical signalling pathways previously implicated in COPD pathogenesis. One of the most striking findings was increased endothelial CXCL-motif chemokine activity and increased signalling amongst monocytes, macrophages, and capillaries, particular from non-classical monocytes to capillary A cells. Therefore, increased signalling between capillaries and myeloid-derived cell populations may be important contributors to endothelial activation in advanced COPD.

In contrast to solely identifying phenotypic shifts amongst endothelial cells with advanced COPD, we observed both phenotypic shifts and population composition changes amongst alveolar macrophages. A subset of macrophages in control lungs was enriched for heat shock proteins while lungs from subjects with advanced COPD were enriched with a cluster expressing metallothionines. Metallothionines are induced by oxidative stress and inflammation and can mediate protective effects by sequestering intracellular metals such as zinc and copper. ^41^ Despite their homeostatic function, little is known about the consequences of chronic metallothionein upregulation in the setting of advanced COPD where redox homeostasis and heavy metal metabolism are disrupted.^42^ For instance, Menkes diseases, a congenital disorder of copper deficiency causes emphysema,^43^ additionally, dysregulated zinc homeostasis can cause impaired phagocytosis and an abnormal inflammatory response in macrophages.^44^ Therefore, understanding metallothionine regulation of zinc and copper in COPD may improve our understanding of disease pathogenesis.

We found enrichment for genetic variants associated with lung function in COPD amongst epithelial and mesenchymal cells but not immune cells. This novel finding suggest that lung resident cells mediate much of the heritability of COPD and suggest the intriguing hypothesis that aberrant immunologic phenotypes in COPD are largely acquired rather than genetically programmed. An effective therapeutic strategy may be targeting epithelial, endothelial, and fibroblast cells in at-risk individuals in early COPD; however, future studies will be necessary to elucidate cell-type specific pathways that underlie COPD heritability.

There are a few limitations that need to be considered. First, COPD is a heterogenous disorder, yet the subjects in this study represented a distinct subtype of COPD patients with advanced disease, requiring transplant, radiographic evidence of emphysema, and who were not actively smoking due to their upcoming lung transplant, and therefore our findings cannot be generalized to all COPD sub-phenotypes or earlier stages of disease. However, our analysis likely reflects the pathologic changes that persist despite smoking cessation. Additionally, while all subjects had radiographic emphysema, we do not know the degree of emphysema within the tissue sample obtained from a given subject. However, given the large number of cells and subjects included in this study, the spectrum of pathologic changes in the lung parenchyma with advanced emphysema were likely captured in our scRNAseq analysis. Other limitations are common to scRNAseq experiments. While transcriptomic changes are informative, they do not always reflect protein concentration, nor do they necessarily impact cellular function. There may also be dissociation bias due variable cellular sensitivities to digestive enzymes and differences in cellular embedding in the extracellular matrix. However, all samples were processed using a previously validated protocol. Finally, findings from the study, particularly the *in silico* CELLECT and connectome analyses, will require further validation.

Overall, scRNAseq identified novel transcriptional profiles of heterogenous populations of AT2, macrophage, and endothelial cell types within the emphysematous lung. Additionally, we demonstrated a key role for lung resident cells in COPD heritability and dysregulation of canonical signalling pathways within the alveolar niche of the emphysematous lung. Together, these findings highlight the complexity of cellular injury, inflammation, and impaired tissue repair that underlies COPD pathogenesis. Future studies evaluating outcomes of specific transcriptional dysregulations identified herein will provide novel insights into mechanisms that contribute to disease.

## Data Availability

Data has been deposited in the Gene Expression Omnibus (GSE136831) and can be explored using our online portal (www.copdcellatlas.com).

## Notes

### Competing Interest Statement

Competing interest: N.K. served as a consultant to Biogen Idec, Boehringer Ingelheim, Third Rock, Pliant, Samumed, NuMedii, Theravance, LifeMax, Three Lake Partners, Optikira over the last 3 years, reports Equity in Pliant and a grant from Veracyte and non-financial support from MiRagen. Has IP on novel biomarkers and therapeutics in IPF licensed to Biotech. All outside the topic of this paper. E.A.A. and S.G.C. are employed by Novartis. M.S. is a co-inventor on a Yale patent application describing the therapeutic utility of MIF020 in lung disease.

### Funding Statement

This work was supported by NIH grants K08HL135402, Flight Attendant Medical Research Institute, and Claude D. Pepper Older Americans Independence Center, Yale School of Medicine to M.S. NIH grants R01HL127349, U01HL145567, U01HL122626, and U54HG008540 to N.K, NHLBI P01 HL114501 and support from the Pulmonary Fibrosis Fund to I.O.R., an unrestricted gift from Three Lake Partners to I.O.R and N.K, and by the German Research Foundation (SCHU 3147/1) to J.C.S.

### Author Declarations

Study protocols were approved by Partners Healthcare Institutional Board Review (IRB Protocol 2011P002419).

